# Integrating Operant and Cognitive Behavioral Economics to Inform Infectious Disease Response: Prevention, Testing, and Vaccination in the COVID-19 Pandemic

**DOI:** 10.1101/2021.01.20.21250195

**Authors:** Justin C. Strickland, Derek D. Reed, Steven R. Hursh, Lindsay P. Schwartz, Rachel N.S. Foster, Brett W. Gelino, Robert S. LeComte, Fernanda S. Oda, Allyson R. Salzer, Tadd D. Schneider, Lauren Dayton, Carl Latkin, Matthew W. Johnson

## Abstract

The role of human behavior to thwart transmission of infectious diseases like COVID-19 is evident. Yet, many areas of psychological and behavioral science are limited in the ability to mobilize to address exponential spread or provide easily translatable findings for policymakers. Here we describe how integrating methods from operant and cognitive approaches to behavioral economics can provide robust policy relevant data. Adapting well validated methods from behavioral economic discounting and demand frameworks, we evaluate in four crowdsourced samples (total N = 1,366) behavioral mechanisms underlying engagement in preventive health behaviors. We find that people are more likely to social distance when specified activities are framed as high risk, that describing delay until testing (rather than delay until results) increases testing likelihood, and that framing vaccine safety in a positive valence improves vaccine acceptance. These findings collectively emphasize the flexibility of methods from diverse areas of behavioral science for informing public health crisis management.

---

The COVID-19 pandemic has spurred important conversations in nearly all sciences [1-3]. While scientists scramble to find ways to address the crisis, a salient role for behavioral science has emerged. Robust evidence documents how human behavior is central to disease spread insofar as reduced travel and social distancing are predictive of lower infection incidence [4, 5] and face mask use effectively mitigates airborne transmission [6, 7]. As of this writing, only remdesivir has been approved by the FDA as an approved safe and effective pharmaceutical. Importantly, remdesivir and other therapeutics are designed for the treatment of existing COVID-19 symptoms rather than for prophylactic prevention, leaving nonpharmaceutical interventions as critical for flattening the curve of transmission [8]. Furthermore, while effective vaccines have been developed and approved under emergency use authorizations, behavioral science remains necessary for ensuring necessary vaccination rates by informing the development of public health programs to counteract factors like vaccine mistrust, skepticism, and apathy [9, 10].

Theoretical commentaries have emphasized the role of understanding human behavior to thwart the transmission of infectious diseases like COVID-19 [11]. Yet, many areas of behavioral science and their accompanying experimental approaches may be limited in the ability to mobilize research to address exponential spread or provide easily translatable findings. For example, methods commonly used in behavioral psychology and the experimental analysis of behavior are limited due to a focus on steady state behavior and within-subject methods [12, 13]. Such designs are hallmarks of rigorous science, to be certain, but these methodological features are sometimes at odds with and constraints on the need for rapid and scalable behavioral solutions. Other behavioral science methods can provide critical data for COVID-19 response, but rely on technical procedures without direct or readily accessible applications that policymakers can act upon [14].

Here, we sought to address these shortcomings by showing how well-validated behavioral economic procedures developed in operant psychology frameworks may be combined with more widely recognized cognitive psychology approaches to provide robust behavioral insights, policy-relevant data, and research methods that are helpful to stakeholders during the COVID-19 pandemic, specifically, but also for infection disease response, more broadly.

Behavioral economics may be defined as an approach to understanding behavior and decision making that integrates behavioral science (commonly psychology) with economic principles [15]. As typically described in the scientific and popular press, this approach emphasizes the contributions of psychology to economics or the *behavior of economics*. Such a behavioral economic tradition often examines how mechanisms described by cognitive psychology can explain systematic deviations from neo-classical economic predictions (e.g., status quo biases; loss aversion) [16-19]. Less often considered, but equally relevant, is the reciprocal integration – the contributions of economics to psychology theory or the *economics of behavior* [20, 21]. Research in this tradition has applied economic principles to understand decision making using methods developed within operant psychology frameworks (e.g., purchasing of competing goods from an economist’s perspective may be the division of operant behavior among competing reinforcers from a behavioral psychologist’s perspective). This approach involves evaluation of behavioral mechanisms including delay discounting (i.e., the devaluation of an outcome by delay), probability discounting (i.e., the devaluation of an outcome by probability/certainty), and behavioral economic demand (i.e., relationship price and consumption that considers this relation may differ across individuals and contexts) to determine how these measurable factors influence choice and behavioral allocation as well as individual difference variables impacting these relationships.

The unique lens by which behavioral economics is used to describes behavior not only provides novel means of interpreting socially important concerns, but also the various facets of the dependent variables generated in these experiments render them especially useful for informing translational public policy [20, 22-24]. For example, operant arrangements can quantify the effective price at which demand for a commodity shifts from inelastic (when a one unit increase in price is met with less than one unit decrease in consumption) to elastic (when a one unit increase in price is met with more than one unit decrease in consumption) for the consumer, while also modeling expected revenue on the part of the supplier. Moreover, comparisons of demand metrics have the potential to determine how imposing different environmental contexts (e.g., availability of reinforcer substitutes, closed economics, framing effects) alters the basic reinforcing value of the commodity for an individual as well as individual factors predictive of that influence. Similarly, discounting procedures can identify the effective delay [25] or probability [26] at which behavior is altered to a given level of performance; for example, the delay associated with, say, a 50% reduction in the value of procuring a COVID-19 test. Such metrics are ripe for modeling policy effects and can provide novel and important behavioral information that is directly relatable to policy makers considering a behavior change program [23, 27, 28].

To date, researchers considering the *behavior of economics* and *economics of behavior* have remained largely independent. We argue that this separation is not of theoretical necessity and that there are many shared interests in cognitive-behavioral factors affecting decision making. A primary goal of this paper is to provide a clear demonstration of how these approaches when integrated can provide scalable behavioral solutions for public health crisis mitigation. To this end, we provide examples that relate to policy designed to reduce transmission and improve treatment within the COVID-19 pandemic. We translated behavioral economic discounting and demand assays widely used and validated in behavioral psychology [29-34] to study 1) engagement in social distancing, 2) cooperation with face mask use, 3) procurement of diagnostic testing, and 4) intentions for vaccination. Within these examples, we evaluate specific experimental manipulations well characterized by cognitive psychology approaches to behavioral economics (e.g., framing effects) to provide clear and translatable implications for public policy design.

## General Methods

### Sampling and Study Overview

This paper summarizes a programmatic series of seven experiments conducted across four samples recruited during the COVID-19 pandemic (Sample 1 N = 133; Sample 2 N = 414; Sample 3 N = 497; Sample 4 N = 322). Sampling occurred asynchronously in March 2020 (Sample 1), May 2020 (Sample 2), July 2020 (Sample 3), and September 2020 (Sample 4). All samples were recruited using crowdsourcing (Amazon Mechanical Turk) with checks used to verify fidelity of responding. One experiment was formally pre-registered (Experiment 7; https://osf.io/56f2z) while others followed standard analyses based on the experimental design. Methods and Results will be presented thematically from prevention behavior to diagnostic testing to vaccination following a general summary of the experimental methods and data cleaning processes.

Across all studies, we required participants to be age 18 or older and have United States residence. Additional attention and validity checks were included for each sample. All studies were reviewed and approved by local Institutional Review Boards (University of Kansas or Johns Hopkins University). Participants reviewed a study cover letter to provide electronic informed consent prior to participation.

### Sample Characteristics and Systematicity Checks

#### Sample 1 (Experiment 1 and 4)

Sample 1 was recruited from 13 March 2020 to 17 March 2020. Participants were required to have a 95% or higher approval rate, 100 or more previously approved tasks, and current United States residence to view and complete the study. Compensation for full study completion was $1 USD. A total of 227 participants completed the full assessment.

Data cleaning consisted of evaluation of behavioral economic tasks for systematic responding as well as evaluation of qualitative responses for English language proficiency and comprehension. Probability discounting task (Experiment 1) and behavioral economic demand tasks (Experiment 4) were evaluated using standardized systematic data checks [35, 36]. A total of 31 participants failed checks on the probability discounting procedure, 18 on the demand procedure, and 44 on both procedures. These results closely corresponded to flagged responses on the qualitative data checks with only one additional participant removed based on inattentive qualitative responses. This resulted in an analyzed sample of 133 participants. The analyzed sample was an average of 39.5 years old (SD = 12.1), 59.8% female, and 80.6% White.

#### Sample 2 (Experiment 5)

Sample 2 was recruited from 13 May 2020 to 10 June 2020. Participants were required to have a 95% or higher approval rate, 100 or more previously approved tasks, and current United States residence to view and complete the study. Compensation for full study completion was $1 USD. A total of 499 participants completed the full assessment.

Data cleaning included evaluation of the diagnostic test delay discounting task (Experiment 5) for reversals (i.e., reversing from stating “No” they would not get a test to “Yes” they would get a test). Any participant with one or more reversal on any task was removed (i.e., 1 task = 7 participants; 2 tasks = 8 participants; 3 tasks = 9 participants; 4 tasks = 61 participants). This resulted in an analyzed sample of 414 participants. The analyzed sample was an average of 32.6 years old (SD = 10.9), 58.3% female, and 71.5% White.

#### Sample 3 (Experiment 2, 3, and 6)

Sample 3 completed the assessment from 4 August 2020 to 12 August 2020. Participants were initially recruited from mTurk in March 2020 as a part of a longitudinal cohort with repeated assessments throughout the COVID-19 pandemic. Participants were required to have a 97% or higher approval rate, more than 100 previously approved tasks, and current United States residence to enroll in the parent study. Data collection for this project occurred in Wave 3 of data collection and included 531 participants who completed the full assessment. Participants were compensated $3.50 USD for completion of this assessment.

Data cleaning included evaluation of social distancing discounting tasks (Experiment 2) and vaccine demand tasks (Experiment 6) for systematic responding. Discounting tasks were evaluated using standardized criteria [35] and demand tasks were evaluated for reversals from a “No” response. A total of 12 participants failed checks on the discounting procedure, 12 on the demand procedure, and 6 on both procedures. An added attention check was included asking about recent use of a fake drug (“oxypentone”) that an additional 6 participants endorsed. This resulted in an analyzed sample of 497 participants. The analyzed sample was an average of 40.0 years old (SD = 11.4), 56.9% female, and 78.7% White.

#### Sample 4 (Experiment 7)

Sample 4 was recruited from 12 September 2020 to 23 September 2020. Participants were required to have a 95% or higher approval rate, 100 or more previously approved tasks, and current United States residence to view and complete the study. Compensation for full study completion was $1 USD. A total of 485 participants completed the full assessment.

Data cleaning included evaluation of the vaccine demand tasks (Experiment 7) for systematic responding. Tasks were evaluated for reversals from a “No” response, which were considered non-systematic (i.e., indicating “No” for intention to get a vaccine and then reversing to “Yes” at a lower efficacy). A total of 163 participants failed checks. This resulted in an analyzed sample of 322 participants. The analyzed sample was an average of 38.8 years old (SD = 11.6), 44.5% female, and 76.7% White.

### Experiment Methods and Data Analysis

All experimental materials are available in the Supplemental Materials. Data were collected via Qualtrics and analyses conducted in *R* Statistical Analysis (see https://osf.io/wdnmx/?view_only=37323f9431aa4c91a0e7209054058dbe for limited datasets and code for primary analyses).

### Social Distancing

Social distancing is a first-line prevention strategy for reducing COVID-19 transmission. In Experiments 1 and 2 we evaluate how the likelihood of engaging in social distancing varies as a function of probabilistic community COVID-19 risk. In Experiment 1, participants completed a probability discounting task evaluating the likelihood of attending a large social gathering given varying probabilities of community risk for a hypothetical disease under varying symptom framing conditions. In Experiment 2, participants completed a probability discounting procedure evaluating the likelihood of engaging in a social activity based on community COVID-19 risk under different risk framing conditions based on a widely disseminated risk assessment infographic from the Texas Medical Association [37].

## Methods

### Experiment 1 (Sample 1)

Participants completed a probability discounting task to evaluate likelihood of attending a large social gathering given the probability of disease risk in the community. The study vignette described a situation involving planned attendance at a large social event. The task included two experimental manipulations related to the symptoms’ description. First, the *symptom type* comprised a within-subject manipulation. A “Mild” version of the task described symptoms including dry cough, fatigue, fever, shortness of breath, and headache. A “Severe” version of the task included these symptoms in addition to difficulty breathing (requiring a medical ventilator). All participants completed these two task manipulations with a randomized order of completion. Second, the *symptom framing* was a between-subject manipulation. Half of participants saw the two task symptom variations with corresponding labels for the symptom severity (e.g., “this group of symptoms is classified as [mild/severe]”) in the “Label” condition (n=69). The other half of participants saw the same symptoms, but with no labels included in the “No Label” condition (n=64). Participants rated their likelihood of attending the social event at varying probabilities that someone in their community was presenting the symptoms described. Participants emitted responses on a visual analog scale (VAS) from 0 (extremely unlikely to attend) to 100 (extremely likely to attend). Symptom probabilities included 0%, 5%, 10%, 25%, 50%, 75%, 90%, 95%, and 100%.

Group discounting data were analyzed and plotted using the hyperbolic discounting equation that includes a non-linear scaling parameter [38]. Individual discounting data were analyzed as area under the curve (AUC) to provide a model free estimation of the impact of symptom probability on the discounting of event attendance [39]. Lower AUC values indicate greater sensitivity to risk (the desirable outcome from a social distancing standpoint). We standardized responses to the 0% likelihood value to isolate the impact of symptom probability from no-risk event attendance. We used an ordinal variation of AUC, here and throughout, to address concerns with normality and disproportional influence of delay or probability steps [40]. AUC values were analyzed using a 2 x 2 mixed ANOVA with the between-subject factor of Label (No Label versus Label) and within-subject factor of Symptom Type (Mild versus Severe). Generalized linear mixed effect models were used to test the likelihood of 100% likelihood of attendance at 0% community transmission risk (a bimodal distribution was observed; therefore, this outcome was dichotomized for analysis).

### Experiment 2 (Sample 3)

Similar to Experiment 1, participants completed a probability discounting task to evaluate likelihood of engaging in a social activity based on community COVID-19 risk. Participants were first asked to select a preferred social activity from a low-to-moderate risk category (i.e., play golf with others, go to a library or museum, or walk in a busy downtown) and from a high risk category (i.e., go to a sports stadium, go to a movie theater, or attend a religious service with 500+ other worshipers). These groupings were based on the risk categorizations made by the Texas Medical Association in June 2020 [37]. Participants then read a vignette describing the opportunity to engage in that activity. A *risk framing* manipulation (between-subject) varied risk categorization with half of participants (N=246) completing task that included labels for the risk severity (e.g., “According to health authorities in your area, this activity is of [Low/High Risk]”) and the other half of participants (N=251) receiving no risk information. The two risk categories were completed in a randomized order. Participants rated their likelihood of going to the social activity at varying probabilities that someone at the activity was displaying COVID-19 symptoms. Participants emitted responses on a VAS from 0 (definitely would not go) to 100 (definitely would not go). Symptom probabilities included 0%, 1%, 5%, 10%, 25%, 50%, 75%, 99%, and 100%.

Group discounting data were analyzed and individual AUC values calculated as in Experiment 1. AUC values were analyzed using a 2 x 2 mixed ANOVA with the between-subject factor of risk Label (No Label versus Label) and within-subject factor of Risk Level (Low-to-Moderate versus High). Generalized linear mixed effect models were used to test the likelihood of 100% likelihood of attendance at 0% community transmission risk.

## Results

### Experiment 1: Framing Effects of Symptom Severity for a Hypothetical Disease

Responding at an aggregate level showed systematic and expected decreases in attendance likelihood based on community symptom risk (Figure 1). Individual AUC values revealed a significant main effect of Symptom Type, *F*_*1*_,_*131*_ = 12.75, *p* < .001, reflecting higher AUC values for the Mild than Severe symptoms, *d*_*z*_ = 0.31. The main effect of Label, *F*_*1*_,_*131*_ = 2.87, *p* = .09, or Symptom Type by Label interaction, *F*_*1*_,_*131*_ = 0.01, *p* = .91, were not statistically significant. Generalized linear mixed effect models testing the likelihood of attendance at 0% community risk also showed no significant differences by Label or Symptom Type, *p* values > .09. *Experiment 2: Framing Effects of Activity Risk for COVID-19*

**Figure 1.**
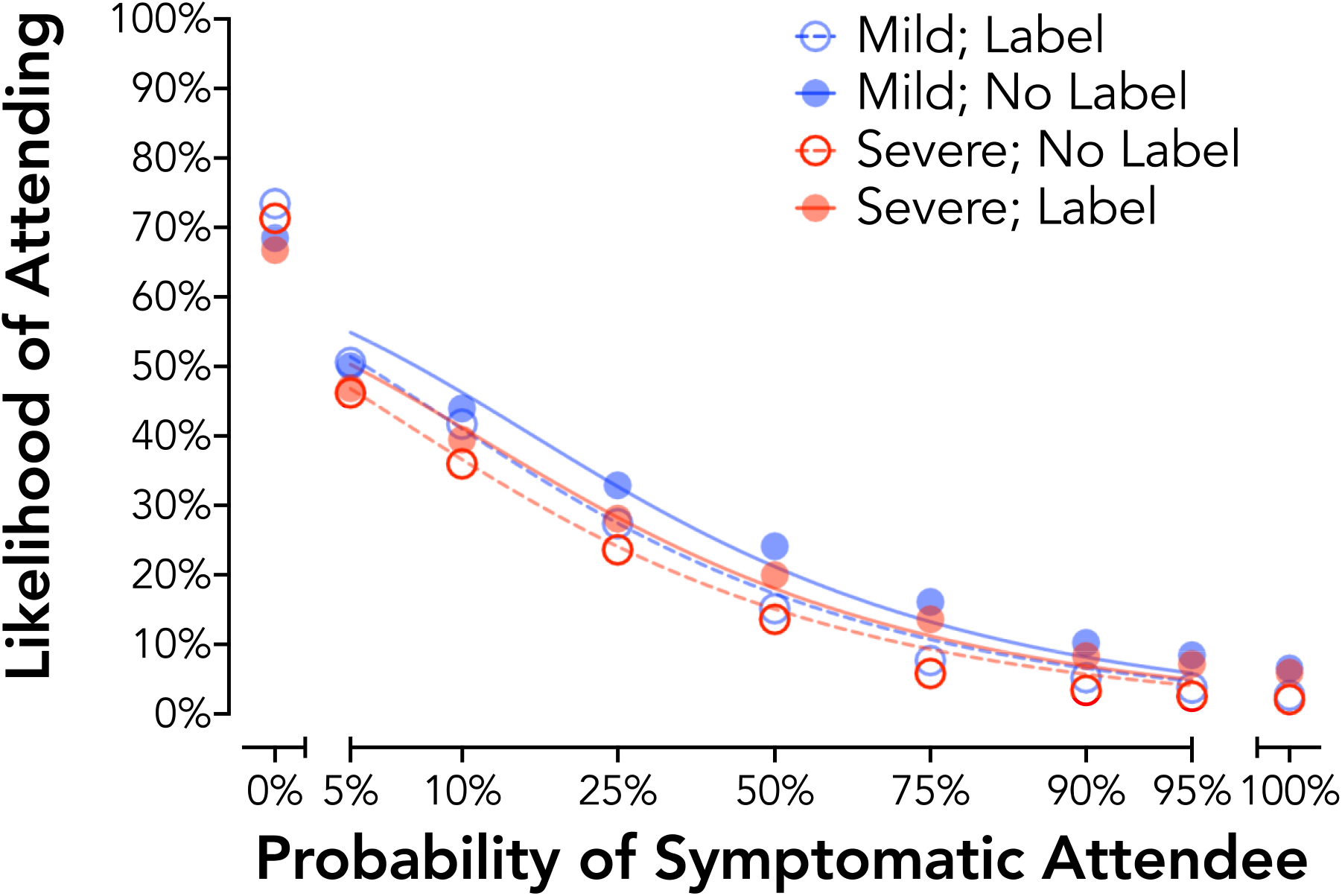
Probability Discounting of Social Event Attendance by Symptom Framing (Experiment 1). Plotted are group discounting curves by severity (mild = blue circles; severe = red circles) and label type (no label = open circles, dotted line; label = closed circles, solid line). Curves are plotted using the hyperbolic discounting equation including a non-linear scaling parameter [38].

Responding at an aggregate level showed systematic and expected decreases in social activity likelihood based on community symptom risk as in Experiment 1 (Figure 2). Standardized AUC values revealed a significant main effect of Risk Level, *F*_*1*_,_*495*_ = 113.62, *p* < .001, and a Risk Level by Risk Framing interaction, *F*_*1*_,_*495*_ = 26.47, *p* < .001. This interaction reflected no significant between-subject effect of Label for the Low Risk activity, *t*_495_ = 0.067, *p* = .95, *d* = −0.01, but in the Label group significantly lower AUC values (i.e., greater sensitivity to risk) for the High Risk activity than the Low Risk activity, *t*_495_ = 2.967, *p* = .003, *d* = −0.27. A significant Risk Level by Risk Framing interaction was also observed at the 0% probability of a symptomatic attendee, *b* = −2.59, *p* < .001. This interaction reflected no differences by Risk Framing in the likelihood of attendance at 0% risk for the Low Risk Activity, OR = 0.84, *p* = .40, but a lower likelihood of attendance for the High Risk Activity for the Label Risk Framing condition, OR = 0.48, *p* < .001.

**Figure 2.**
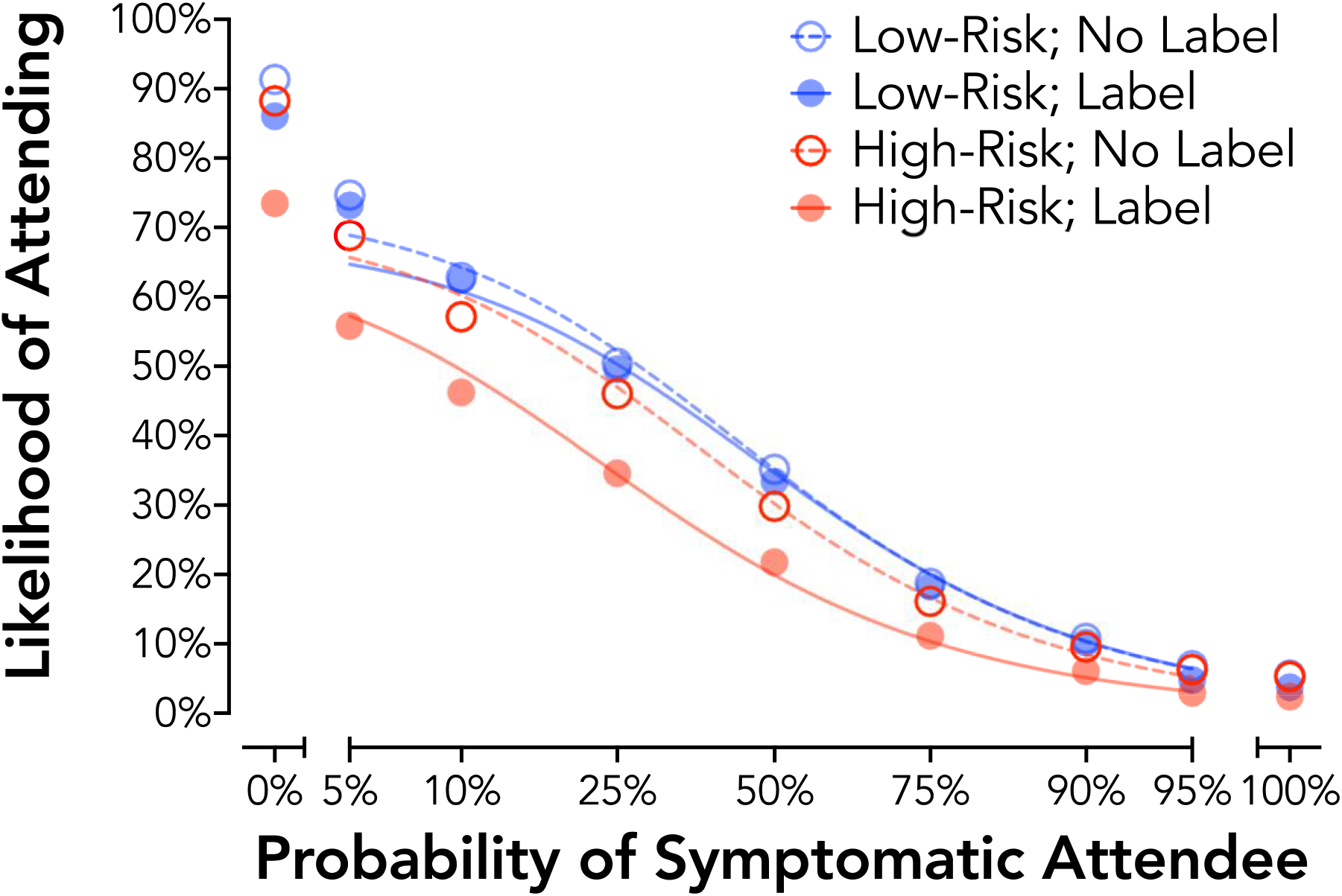
Probability Discounting of Social Event Attendance by Risk Framing (Experiment 2). Plotted are group discounting curves by risk (low risk activity = blue circles; high risk activity = red circles) and label type (no label = open circles, dotted line; label = closed circles, solid line). Curves are plotted using the hyperbolic discounting equation including a non-linear scaling parameter [38].

## Discussion

These findings collectively show that social distancing can be effectively modeled using probability discounting procedures. Social activity was systematically devalued by likelihood of non-specific (Experiment 1) and COVID-19 specific (Experiment 2) community disease risk. Importantly, we found that a framing manipulation modeled after popular public health messaging targeting social distancing increased sensitivity to risk likelihood for a high-risk activity while not appreciable changing behavior for a low-risk activity. This is important given the potential for risk framing to have an untoward effect of reducing risk sensitivity for low-risk settings when presented in these kinds of behavioral contrasts (i.e., boomerang effects) [41].

### Facemask Use

Consistent face mask use in social interactions is one of the most widely recommended and effective means of reducing COVID-19 transmission [6, 7]. Despite this effectiveness, the use of face masks remains controversial and underutilized [42]. Evaluating individual and contextual factors that influence face mask use may help to identify areas for intervention – either at a population level (stemming from between-person differences) or at a contextual level (stemming from within-person differences).

In Experiment 3 (Sample 3) we evaluated the role of social factors in determining face mask use. The notion that face mask use may relate to social context is reasonable given that one of the primary benefits of mask use is prevention of transmission to others. With respect to behavioral economic theory, social discounting is a well-described behavioral mechanism by which the value of an outcome is devalued by the “social distance” or subjective “closeness” of a person to the participant (e.g., a co-worker would be more socially distant than a sibling or parent). Empirical work on social discounting finds that the value of an outcome is hyperbolically devalued by social distance in a way that is mechanistically similar to delay and probability discounting [43-45]. Participants in Experiment 3 completed a traditional social discounting task using monetary consequences and a novel social discounting task with face mask use as the consequence.

## Methods

### Experiment 3 (Sample 3)

Participants completed social discounting tasks evaluating responding for face mask and monetary outcomes [45]. Prior to completion of the social discounting tasks, participants were asked to think of the 100 people closest to them with 1 being the dearest friend or relative in the world and 100 being a mere acquaintance. Participants were then asked to record their relation to people at numbers 5, 10, 20, 50, and 100 on this list, all of which were people instructed to be someone they did not live with or see in the past month. This information was included in the response options to personalize responding. In the face mask version of the task, participants were instructed to report their likelihood of wearing a face mask when interacting with people at each of these social distances using a 100-point VAS. Three conditions were presented including 1) when the participant was COVID-19 asymptomatic, 2) COVID-19 symptomatic without a positive test, and 3) COVID-19 symptomatic with a positive test. These conditions were presented in that order as a means to model the progression that a person may experience in decision-making (i.e., asymptomatic to positive). Participants also completed a social discounting task for money based on prior methods [43]. In this task, participants were asked to select between receiving an amount of money for themselves alone or $75 USD for the N person on the list. Participants completed this task for the 5, 10, 20, 50, and 100 on their social list. Participants were also asked about responding for a stranger in each task.

Non-systematic data were removed for Experiment 3 specific to Experiment 3 rather than for all experiments using Sample 3 data. This was done given the unorthodox nature of the social discounting task and unusual pattern of response consistently observed (see below), meaning that non-systematic responding was less likely to represent non-specific responding. Of the 497 participants in the Sample 3 analyzed set, 45 showed non-systematic responding on any of the social discounting tasks for face masks for an Experiment 3 analyzed sample of 452 participants. Group discounting data were analyzed as in Experiment 1. Generalized linear mixed effect models were used to test the likelihood of using a face mask across all social distances as a function of condition (asymptomatic, symptomatic no test, symptomatic test).

## Results

Prior to data collection, we expected that the likelihood of using a face mask would be devalued by social distance such that the greater the social distance, the lower likelihood of mask use. Surprisingly, an opposite pattern of behavior was observed – participants reported *greater* likelihood of mask use with increasing social distance in an orderly fashion (Figure 3; bottom right). These data suggesting a systematic discounting pattern, but only if the discounted outcome was social interaction *without* a face mask rather than use of a face mask. In fact, when recoding responses to measure likelihood of interaction without a face mask, a systematic discounting pattern was observed with the likelihood of interacting without a mask discounted hyperbolically by social distance (Figure 3; bottom left). Critically, responding on the monetary social discounting task followed an expected and typical pattern in which the amount of money foregone to a social partner was systematically and hyperbolically discounted by increasing social distance (Figure 3 top panel).

**Figure 3.**
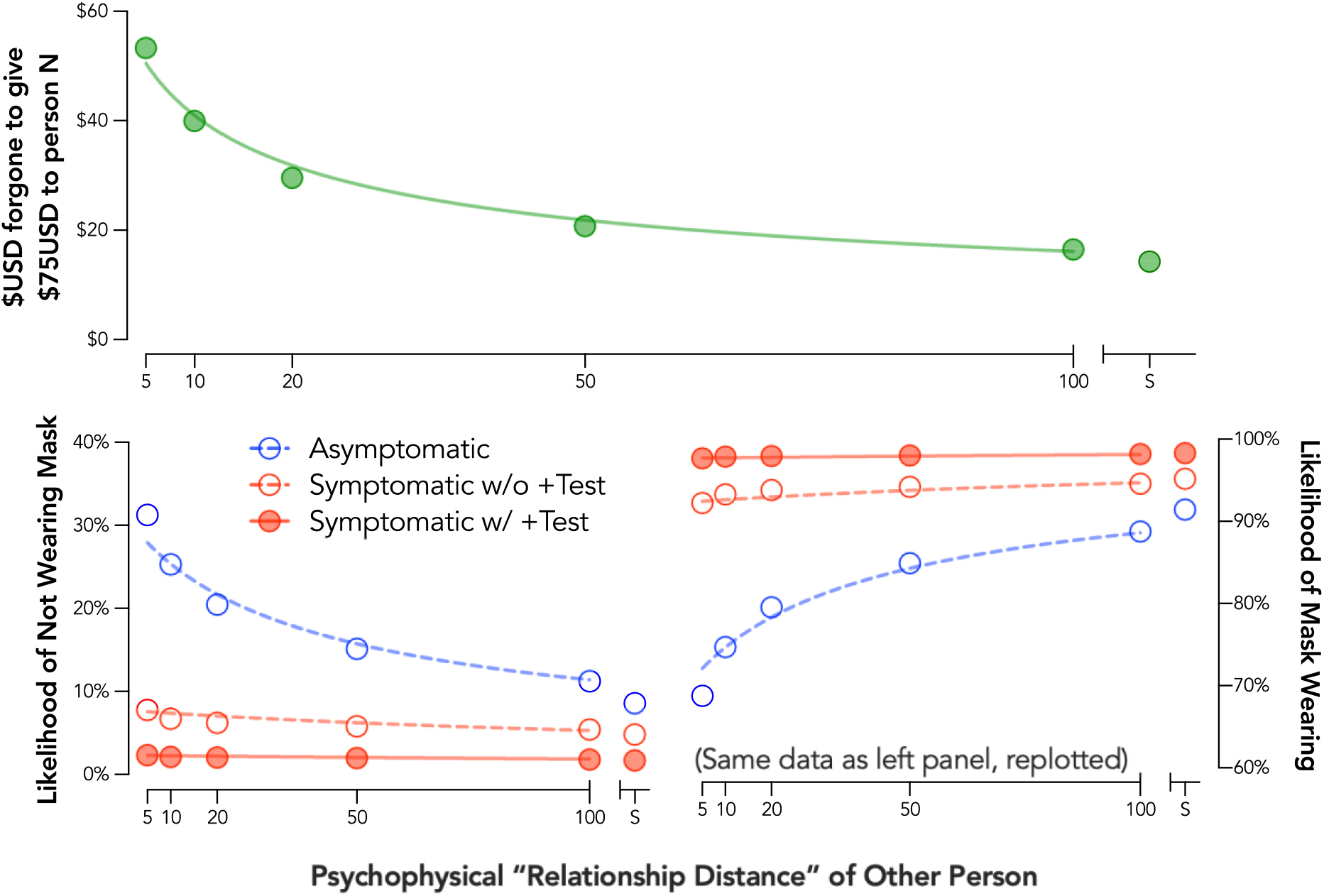
Social Discounting for Face Mask Use and Monetary Outcomes. Plotted are group discounting curves for money (top panel) and face mask use (bottom panels). Three face mask use conditions are presented: asymptomatic (red open circles, dotted line), was symptomatic without a COVID-19 test (red open circles, dotted line), and 3) was symptomatic with a positive COVID-19 test (red closed circles, solid line). Curves are plotted using the hyperbolic discounting equation including a non-linear scaling parameter [38]. S = stranger.

A clear effect of symptomatic status was also observed such that face mask use was more likely (i.e., likelihood of forging a mask was lower) when a participant was symptomatic whether with or without a positive test (Figure 3). This pattern of response was attributable, in part, to an increase in the proportion of participants reporting they would wear a mask when interacting with any social partner in the asymptomatic condition (44.9%) to the symptomatic condition (78.8%, OR = 32.1, *p* < .001) and positive test condition (94.0%, OR = 604.1, *p* < .001).

## Discussion

These findings collectively show that face mask use is sensitive to social factors well described by social discounting and behavioral economic procedures. Our assumption based on existing social discounting research was that one would be more likely to wear a mask with those who are interpersonally close given concern about infecting those they care about. However, it appears that the increased value of maskless interaction with those one is interpersonally closer to may overshadow any additional concern for that person’s safety. Unsettlingly, these findings suggest that the value of mask-less social interaction is the more salient factor considered when deciding whether to use or not use a mask based on social relations to others. Relevant to note is that participants were asked to respond to these questions for people that they had not seen in the past month and did not live with. Therefore, these findings cannot be accounted for by responding based on people who they person has already had recent close and mask-less contact with. Although additional work is needed to tease apart specific factors contributing to these findings, these data are in line with other literature emphasizing the value of facial expression for emotional and social interaction [46, 47]. This unexpected, but systematic finding emphasizes that efforts to convey the relevance of mask use even when interacting with those you know well is warranted when promoting consistent face mask use.

### Diagnostic Testing

COVID-19 testing is key for identifying infection status to prevent future transmission as well as to inform contact tracing. However, difficulties in obtaining testing and subsequent delays related to receiving results have been a noted criticism of COVID-19 efforts. Experiments 4 and 5 were designed to evaluate these testing decision-making processes. Participants in Experiment 4 completed a task evaluating demand for a diagnostic test following possible exposure to a hypothetical disease with the symptoms of cough, fever, and shortness of breath. Experiment 5 was designed to build on these findings with direct applications to COVID-19 by evaluating the impact of cost and delay for COVID-19 diagnostic testing.

## Methods

### Experiment 4 (Sample 1)

Participants completed a hypothetical purchase task procedure to evaluate behavioral economic demand for a diagnostic testing kit for a hypothetical disease. Specifically, participants read a vignette indicating that they had attended a social event with over 200 people and one week later developed symptoms including cough, fever, and shortness of breath. Participants were also instructed that one other person in their county had developed an infection, that a nearby hospital or clinic had a testing kit, but that there were no others in the area, that this kit was approved by the Centers for Disease Control and Prevention (CDC), and that they had their typical income and savings available when making these decisions. Participants were asked to report the likelihood of purchasing a testing kit given a series of out-of-pocket costs ($0 [free], $1, $5, $10, $20, $30, $40, $50, $75, $100, $150, $200, $500, $1,000, $2,000, and $5,000/kit). Participants emitted responses on a VAS from 0 (extremely unlikely to get tested) to 100 (extremely likely to get tested).

Individual demand data were evaluated using curve observed values including demand intensity (reported likelihood of consumption at zero price), O_max_ (individual maximum expected expenditure), P_max_ (price at individual maximum expected expenditure), and breakpoint (BP1; last price at which any likelihood of consumption occurred). Demand intensity was analyzed as a dichotomized variable of 100% likelihood of getting a testing kit at zero price versus < 100% likelihood given the observation of clustering (i.e., 81.2% of participants indicating they would definitely get tested if free). O_max_ and P_max_ were also square-root transformed prior to analysis to reduce variable skew. Bivariate correlations were conducted as Spearman correlations between four preventive health behaviors (i.e., hand washing, face touching, social distancing, and avoiding large groups; recorded on a 1 to 5 scale of never to all the time) and demand measures. Group mean demand curve was also fit using the exponential demand equation [48] to evaluate the analytical P_max_ value reflecting the point at which a one-log unit increase in price is met by a one log-unit decrease in consumption [49].

### Experiment 5 (Sample 2)

Participants completed a delay discounting procedure in which decisions to obtain testing were assessed across systematically varied delays (15 minutes to 28 days). We evaluated two within-subject manipulations in a factorial design. First, *cost* was manipulated with a test as Free or $125 in out-of-pocket expenses (based on the distribution of out-of-pocket costs for COVID testing at the time of the study). Second, *delay framing* was manipulated with one set of tasks evaluating delay to receiving a test kit with immediate results and the other set evaluating delay to receiving results after an immediate test. Delays were held consistent across these two delay types such that the only stated differences were in the framing of the delay. Participants completed the testing delay condition prior to the results delay condition with price randomized within these two conditions. Participants were asked if they would get a testing kit given a series of delays (15 min, 60 min, 1 day, 2 days, 3 days, 5 days, 7 days, 14 days, and 28 days). Response options for Experiments 5 as well as Experiments 6, and 7 were simplified as dichotomous yes/no choices rather than the VAS used in prior tasks. This design feature was selected to streamline responding and better model actual decision-making in which decisions are a discrete yes or no choice.

Group data were modeled as in Experiment 1. Maximum delay for each condition was used as a within-subject measure and calculated as the individual median value between last accepted and first rejected delay. Higher maximum delay values are indicative of acceptance of longer imposed delays. Maximum delay values were analyzed using a 2 x 2 repeated measures ANOVA with the within-subject factors of risk Price (Free versus $125) and Delay Framing (Delay to Test versus Delay to Result).

## Results

### Experiment 4: Sensitivity of Diagnostic Testing to Cost

Demand for a diagnostic test systematically decreased with increases in cost with the exponential demand model describing aggregate demand well (Figure 4; *R*^*2*^ = 0.99). The group average demand curve indicated an analytical P_max_ value of $207 indicating the price at which the demand curve shifts from inelastic or sub-proportional sensitivity of consumption to price to elastic or super-proportional sensitivity of consumption to price. Evaluation of individual demand curve values indicated that greater engagement in hand washing was significantly associated with greater demand when free, *r* = .23, *p* = .009, and maximum expenditure for a test, *r* = .21, *p* = .015. Similarly, greater avoidance of face touching was significantly associated with greater demand when free, *r* = .23, *p* = .009, and maximum expenditure for a test, *r* = .19, *p* = .027.

**Figure 4.**
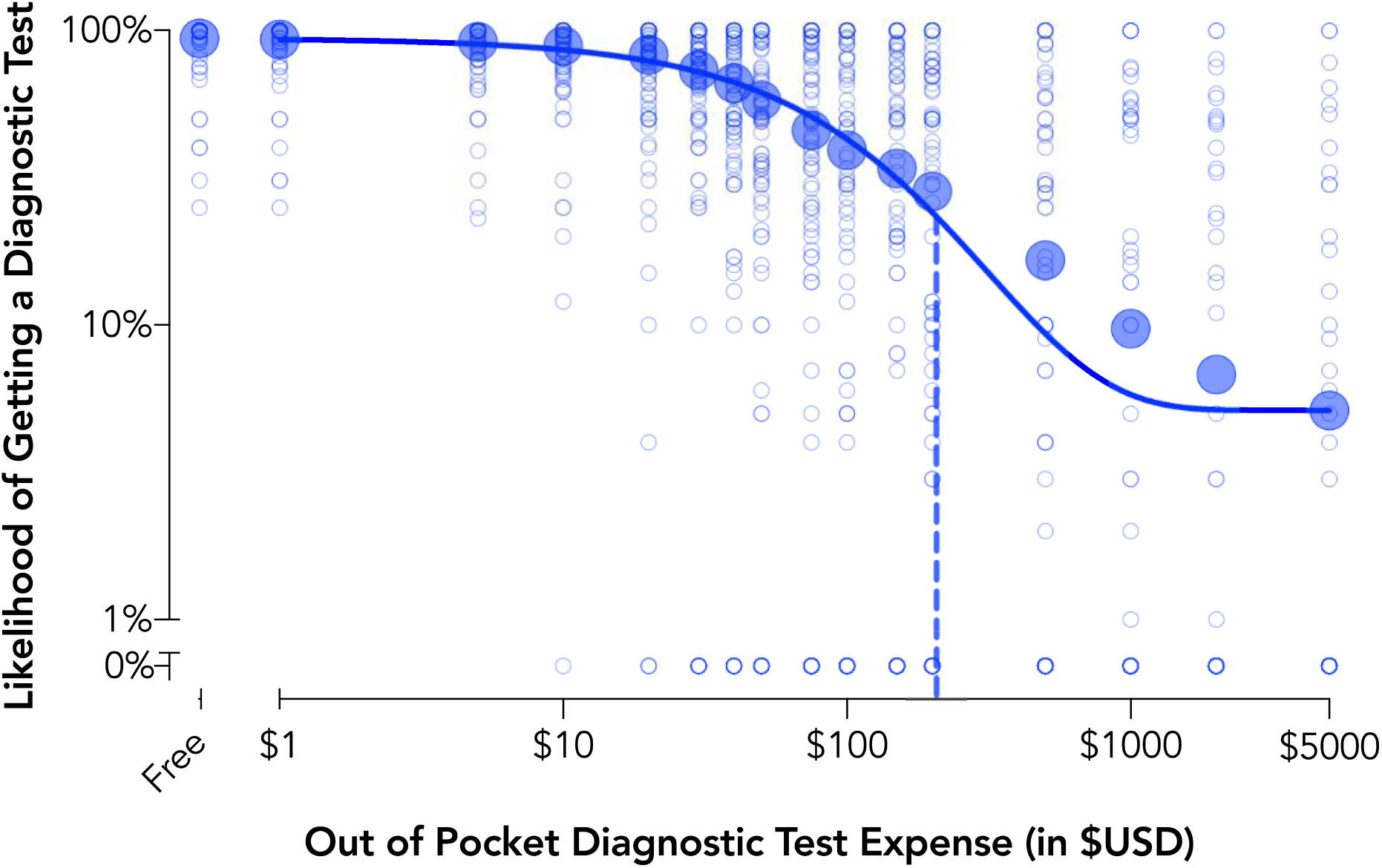
Behavioral Economic Demand for Diagnostic Testing (Experiment 4). Plotted are group mean data and individual data points for behavioral economic demand of diagnostic testing recorded on the hypothetical purchase task. Demand curve data are plotted using the exponential demand function [48]. The dotted line is the price representing shifts from inelastic or price insensitive to elasticity or price sensitive demand (P_max_).

### Experiment 5: Delay to Test versus Delay to Result in COVID-19 Testing

Assessment of aggregate discounting curves showed systematic reductions in testing intentions with increases in delay for each condition (Figure 5). Tests of individual subject values (crossover delay from yes to no testing intention) found significant mains effects of Price, *F*_*1*_,_*413*_ = 523.8, *p* < .001, and Delay Framing, *F*_*1*_,_*413*_ = 23.1, *p* < .001, and a Price x Delay Framing interaction, *F*_*1*_,_*413*_ = 30.4, *p* < .001. Evaluation of this interaction indicated that longer delays were tolerated when the delay was to receive a test rather than receive results when tests were free, *t*_*413*_ = 5.64, *p* < .001, *d*_*z*_ = 0.28, but that there were no significant differences by framing when tests had out-of-pocket costs, *t*_*413*_ = 0.73, *p* = .47, *d*_*z*_ = 0.04.

**Figure 5.**
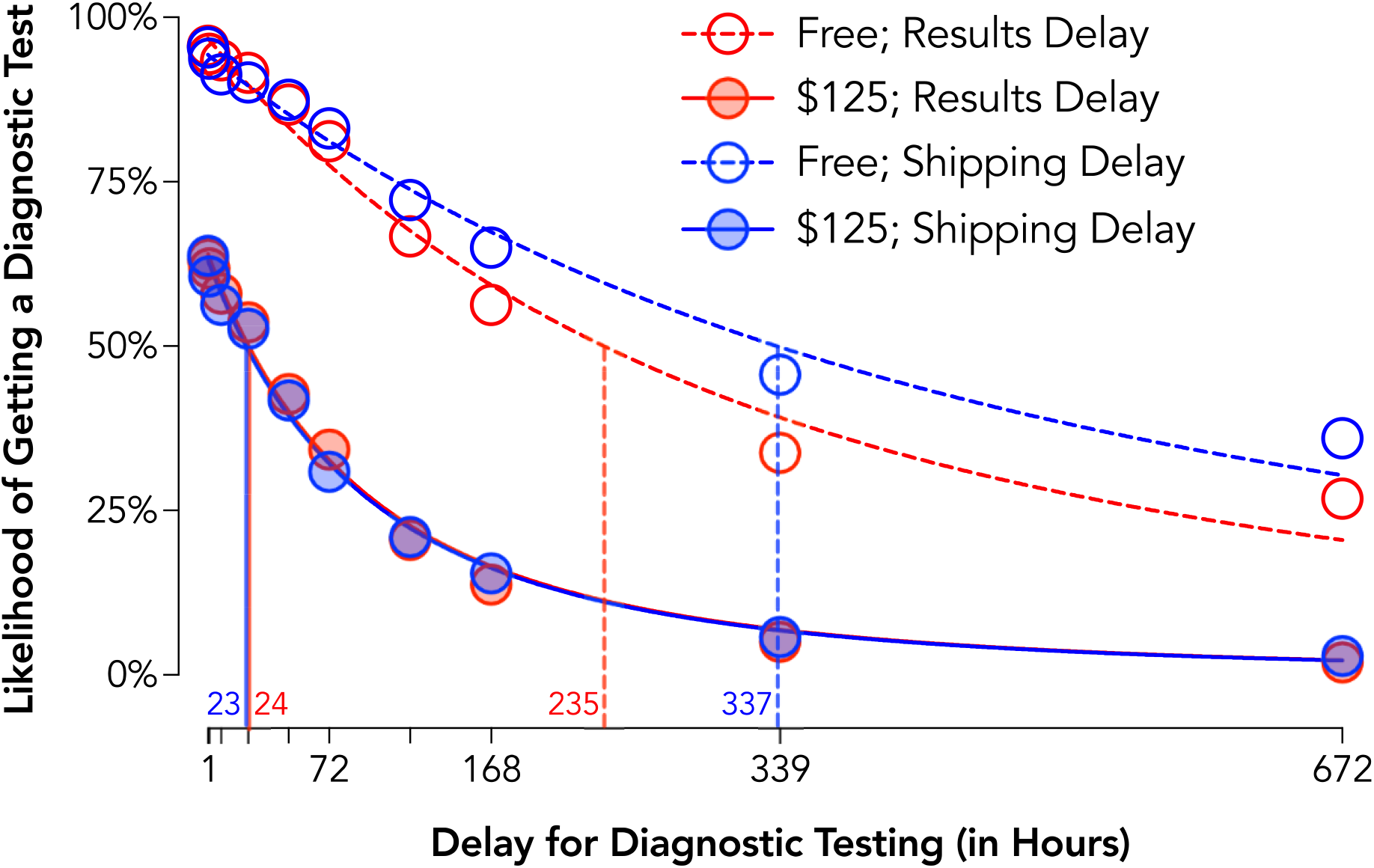
Delay Discounting of COVID-19 Diagnostic Testing by Delay Type and Cost (Experiment 5). Plotted are group discounting curves by delay type (delay to receiving a test with immediate feedback = red circles; delay to receiving results from an immediate test = blue circles) and cost (Free = open circles, dotted lines; $125; closed circles, solid lines). Curves are plotted using the hyperbolic discounting equation including a non-linear scaling parameter [38]. Vertical lines are estimated ED50 or the delay at which half of the population is likely to procure a test.

These findings are highlighted in differences for ED50 values (Figure 5 vertical lines) summarizing the delay at which half the population is likely to procure a test. Specifically, when participants had to pay $125 for test, ED50s of approximately 1 day (23 and 24 hours) were observed in both framing conditions. In contrast, when Free, the ED50 was 4.25 days longer (102 hours) for the shipping delay than results delay condition with lower sensitivity to delay in the shipping delay condition.

## Discussion

Experiments 4 and 5 collectively show that diagnostic testing is sensitive to factors including testing cost and delay. Importantly, these data emphasize how delays imposed on receiving test results may exert a particularly strong impact for discouraging testing, emphasizing how rapid testing may improve testing rates even if a delay is imposed on getting the test. That responding was more sensitive to delayed results than delayed testing is possibly explained by a dominant response (i.e., getting a test) outcome (i.e., receiving a result) contingency at play and how delays for this response-outcome contingency are exaggerated under a delayed results scenario.

### Vaccination Intentions

Recent emergency authorization of and attempts at distribution of vaccines for COVID-19 have highlighted challenges related to vaccine skepticism and the role of behavior change and motivation as key steps for encouraging vaccine uptake. Experiment 6 evaluated demand for both a COVID-19 vaccine and an influenza vaccine based on the efficacy of those vaccines. We used an experimental vignette in which the participant was at a health care provider and could “bundle” an additional vaccine with the one they were already receiving. Experiment 7 evaluated a choice framing condition in which COVID-19 vaccination safety was framed positively or negatively.

## Methods

### Experiment 6 (Sample 3)

Participants read vignettes describing a scenario in which approved influenza and COVID-19 vaccines were available. The instructions indicated these vaccines would be the only ones available, that they would be free of cost, would have to be administered now, and were approved by the FDA. Scenarios were presented to model going to a healthcare provider for one vaccine and having an option to bundle another vaccine at that visit. Participants responded across a series of efficacies defined as percentage reduction in influenza/COVID-19 symptom risk (100% to 0% effective in 10% increments). Participants were randomized to complete different *choice framing* conditions (between-subject). In an opt-in condition, the response option was preselected as “No” and participants were required to change the selection to “Yes” if they wanted the vaccine (n = 245). In an opt-out condition, the response option was preselected at “Yes” and participants were required to change the selection to “No” if they wanted the vaccine (n = 252). All participants also completed a version in which no response was preselected and were randomized to complete this before or after the choice framed condition.

Group data were modeled using demand methods as in Experiment 4. Individual values for minimum required efficacy for each vaccine task were calculated as the individual median value between last accepted and first rejected vaccine efficacy. Individuals who rejected the vaccine at all values were assigned a value of 100 and those accepting at all values were assigned a value of 0. Higher minimum required efficacy values are indicative of a need for higher vaccine efficacy for vaccine intention. Minimum required efficacy were first analyzed using a 2 x 2 x 2 mixed ANOVA with the within-subject factors of risk Vaccine Type (COVID-19 and Influenza), Response Type (Default versus No Default) and the between-subject factor of Framing Condition (Opt-In versus Opt-Out). A secondary analysis was conducted with only the first framing condition completed as a 2 x 3 mixed ANOVA with the within-subject factor of Vaccine Type (COVID-19 versus Influenza) and Response Condition (No Default, Opt-In, and Opt-Out).

### Experiment 7 (Sample 4)

Experiment 7 was conducted with a preregistration (https://osf.io/56f2z). Participants completed demand tasks in which we varied *development timeline* (within-subject) as either a 7-month (for late October 2020 delivery) or 12-month (for late March 2021 delivery) process to model scenarios presented in news media at the time of data collection (September 2020). Participants were randomized to a *safety framing* condition (between-subject) in which safety was described using a positive framing (“95% of the scientific community declares the vaccine safe”; n = 161) or a negative framing (“5% of the scientific community declares the vaccine unsafe”; n = 161). Assignment was stratified based on endorsement of receiving a flu vaccine in the past three years to ensure balance in general vaccination behavior between the two conditions.

Group data were modeled using demand methods as in Experiment 4 and individual required minimum efficacy calculated as in Experiment 6. Minimum required efficacy data were first analyzed using a 2 x 2 mixed ANOVA with the within-subject factors of risk Development Timeline (7-month versus 12-month) and the between-subject factor of Framing Condition (Positive versus Negative Framing). A secondary analysis was conducted with only the first task completed as the same 2 x 2 ANOVA with Development Timeline as a between-subjects factor. A sensitivity analyses was also conducted including the covariates of age, gender, and conservativism (Social and Economic Conservatism Scale) [50]. This analysis used a linear mixed effect model including these covariates, the fixed effects of Development Timeline and Framing Condition, and a random intercept term. A deviation from the preregistered analysis plan was made for this sensitivity analyses because education was not collected in the survey, and therefore, not available to include as a covariate.

## Results

### Experiment 6: Opt-In and Opt-Out Procedures for COVID-19 and Influenza Vaccine Bundles

Aggregate demand curves showed systematic decreases in demand for a vaccine with decreases in efficacy (Figure 6). The exponential demand model described aggregate demand well across each demand curve and allowed for estimation of vaccine coverage at a critical threshold (e.g., 70% coverage) [51]. Analysis of individual cross-over efficacies (i.e., the efficacy at which a participant went from intending to not intending vaccination) revealed a significant main effect of Vaccine Type, *F*_1,495_ = 39.3, *p* < .001, reflecting vaccine acceptance at lower efficacies for a COVID-19 vaccine than an influenza vaccine. Main effects and interactions involving the framing condition were not significant, *p* > .10.

**Figure 6.**
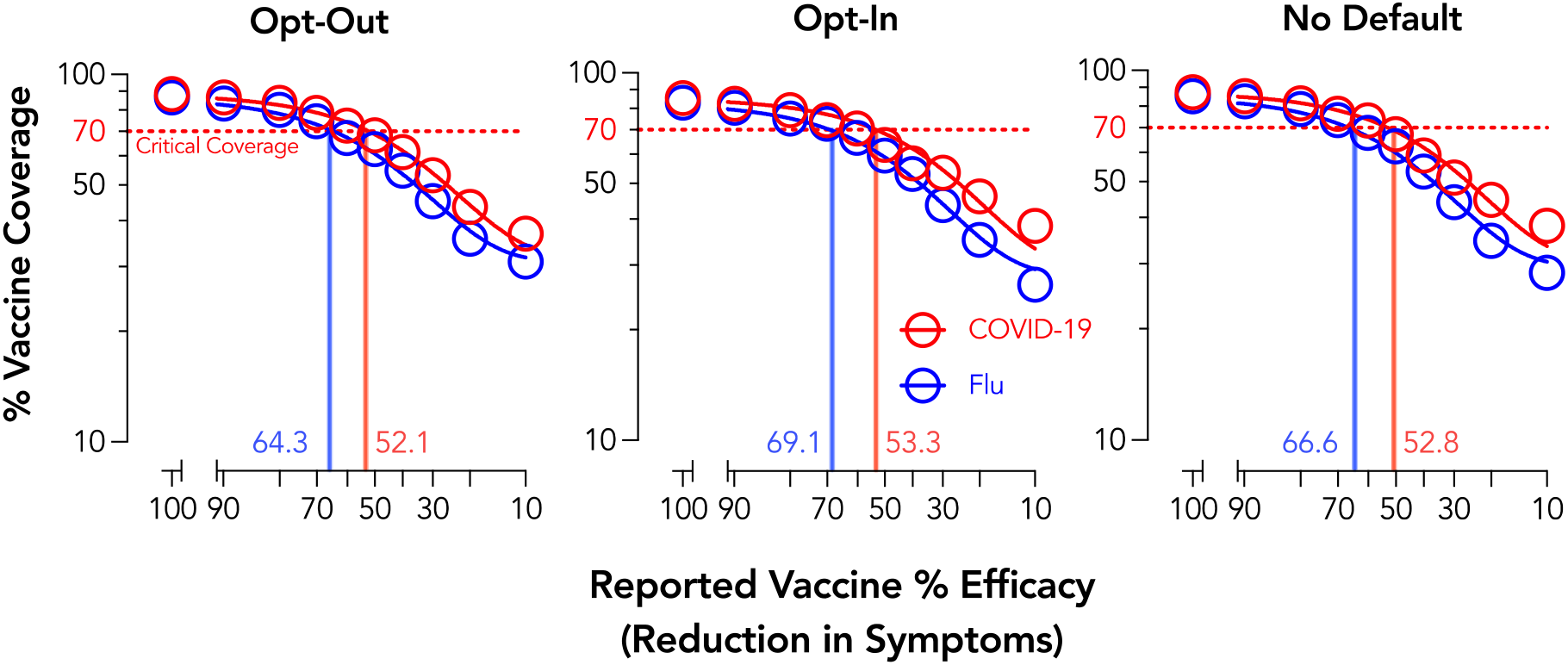
Vaccine Acceptance by Efficacy, Type, and Choice Framing. Plotted are group discounting curves by vaccine type (COVID-19 = red; flu = blue). Demand curve data are plotted using the exponential demand function [48]. Vertical lines plot the efficacy needed to reach a critical coverage of 70%.

### Experiment 7: Development Timeline and Safety Framing for COVID-19 Vaccination

Aggregate demand curves showed systematic decreases in demand for a vaccine with decreases in efficacy across each condition (Figure 7). At an individual level, significant main effects of Development Timeline, *F*_1,320_ = 9.04, *p* = .003, and Safety Framing, *F*_1,320_ = 14.86, *p* < .001, were observed. These effects reflected acceptance of less effective vaccines under a positive framing condition, *d* = 0.33, and when developed for longer, *d*_*z*_ = 0.22. Controlling for age, gender, and political conservatism did not change the results of these findings. Evaluation of these effects with only the first development time completed (i.e., a purely between-subject design) indicated a similar main effect of Safety Framing, *F*_1,318_ = 7.32, *p* = .007, but found that the Development Timeline effect was no longer significant, *F*_1,318_ = 2.31, *p* = .13. Post-hoc analysis of this possible carryover effect indicated that the Development Timeline effect was statistically significant for participants that completed the 12-month condition first, *t*_*160*_ = 4.77, *p* < .001, *d*_*z*_ = 0.38, but not the 7-month condition first, *t*_*160*_ = 0.73, *p* = .47, *d*_*z*_ = 0.06.

**Figure 7.**
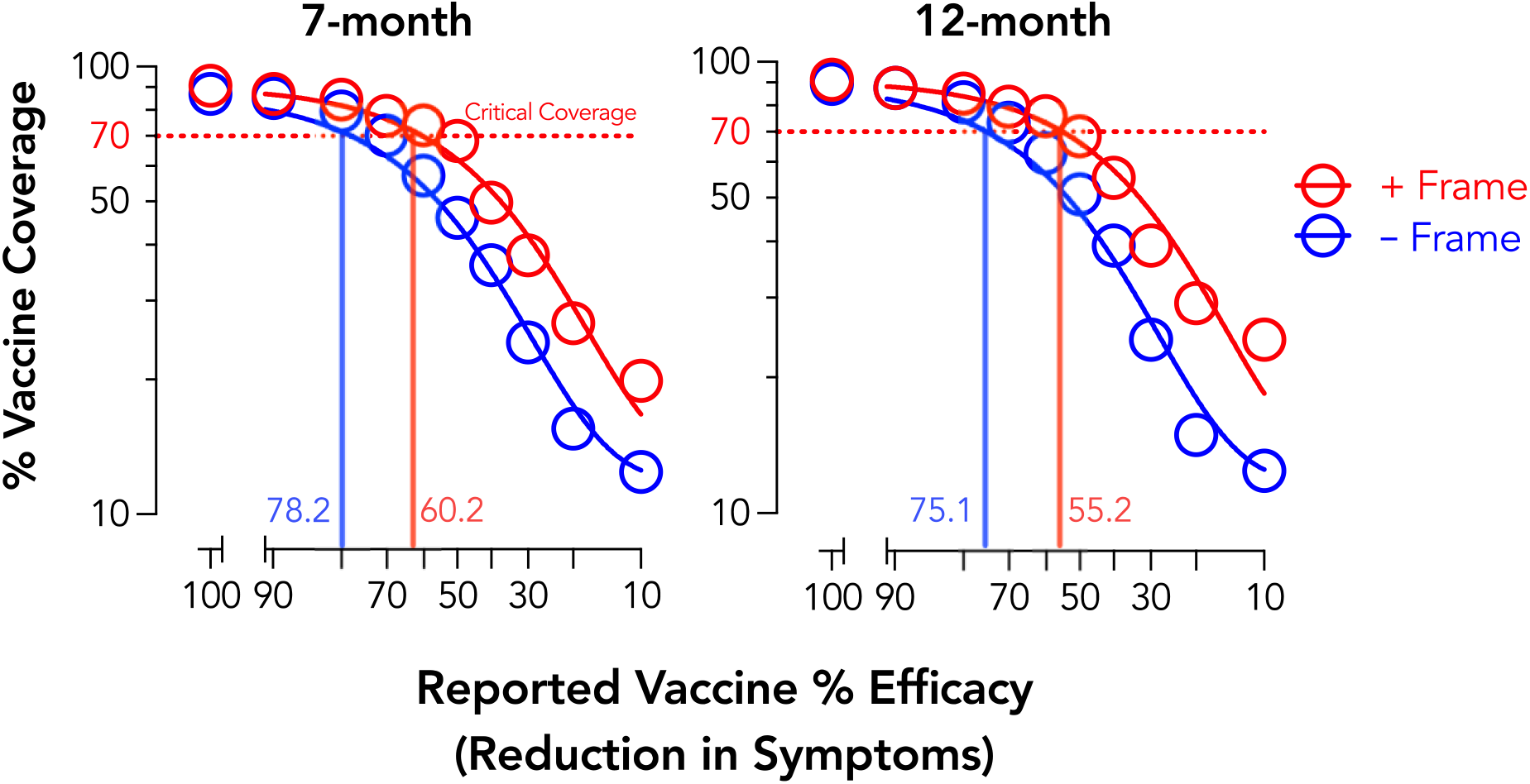
COVID-19 Vaccine Acceptance by Development Timeline and Safety Framing. Plotted are group discounting curves by safety framing (positive = red; negative = blue). Demand curve data are plotted using the exponential demand function [48].

## Discussion

Experiments 6 and 7 found that vaccination intentions were systematically related to efficacy, both for a COVID-19 vaccine and an influenza vaccine. Experiment 6 did not reveal a significant effect of choice framing, which is possibly due to the online setting and limitations of modeling these kinds of opt-in/opt-out procedures. A substantive framing effect for vaccine safety, however, was observed such that intentions were lower under a negative than positive framing. These findings are relevant in that news sources – even when presenting the same data – may focus on either positive (% of scientists approve) or negative (% of scientist disapprove) framings when conveying this information to its readership or viewership [for similar issues in climate change messaging see 52]. The current findings show how such framings could adversely impact the likelihood of obtaining a vaccine and ways in which public health messaging should be optimized to avoid such biases.

### General Discussion

The COVID-19 pandemic has emphasized how behavioral science is critical to informing public health crisis management. In the current study, we sought to determine how behavioral economic approaches developed from cognitive psychology and operant behavioral psychology traditions can be integrated to address existing and emerging issues in public health – doing so in a rapid and scalable manner. Adapting well validated methods from behavioral economic discounting and demand frameworks, we evaluated behavioral mechanisms contributing to the engagement in preventive health behaviors relevant to infectious disease transmission, namely those associated with the COVID-19 pandemic. We also evaluated how framing manipulations can alter decision-making in ways relevant to public health and policy implementation. These findings collectively emphasize how merging behavioral economics methods can rapidly generate empirical data to inform public health crisis management while retaining a strength informed by foundational conceptual frameworks for health behavior change.

The present study advances behavioral science in several ways with each contribution emphasizing its ability to address critical and acute public health crises that may not be amenable to prototypical experimental methods. First, this study translates operant discounting and demand methods to simulate decision-making in an uncommon context for which an individual has no direct experience. The COVID-19 pandemic is a public health crisis, the likes of which have never been experienced by anyone alive today. Although hypothetical discounting and demand tasks are presumed to reflect verbal behavior shaped by histories of consequences in similar choice contexts [53, 54], some decisions lack formal similarity with actual experience. Decisions regarding social isolation, diagnostic testing, or vaccinations for an infectious disease pandemic are relatively novel and require participants to consider generalized decision-making repertoires, such as deciding to take precautions in avoiding individuals with the common cold or influenza virus. A small, but growing, literature suggests that these kinds of tests of novel or as-yet-unexperienced contexts can nonetheless significantly relate to real-world behavior of interest. For example, in the public health domain, studies on sexual discounting relate to HIV-risk behavior [55, 56] and simulated purchasing of a novel fake ID relate to experienced negative alcohol outcomes in underage drinkers [57]. Moreover, there is evidence that tasks such as hypothetical sexual discounting [58] or hypothetical purchase tasks for drugs [59, 60] significantly predict domain-specific outcomes or behavior beyond general monetary discounting or demand for common commodities. The current study adds to this literature while extending to the study of infectious disease and pandemic response.

Second, the data provided by this approach permits safe modeling of potential public health policies. Hursh [20] previously outlined proposed strategies for how behavioral economics can inform health policy, suggesting the quantification of commodity valuation in behavioral economic analyses lend well to informing policy-making. Specifically, experimental research permits controlled and accurate measurement, which may lend new behavioral insights into econometric analyses of market behavior. This information may then inform the creation of experimental model projects to measure scalable policy-level interventions at the community-level. Successful results thereby lead to policy formation, implementation, and evaluation; if there are failures, such results form a feedback loop wherein behavioral scientists can seek to modify procedures and policies to re-evaluate such effects. Related work in psychology and related fields has harnessed hypothetical discounting and demand techniques to provide novel lenses by which to view population-level effects for hard-to-study behavioral questions – from a direct operant perspective, at least – such as tornado warnings [61], incremental cigarette taxation [27], texting-while-driving interventions [62], and happy-hour pricing for alcohol [63];such findings speak directly to potential population-level decisions and have an added benefit of providing accurate quantitative markers for policy development and targets [20, 22, 23].

Finally, this study has consequences for understanding behavioral phenomena directly concerning the spread of COVID-19: social distancing, face mask use, testing procurement, and vaccination intentions. Across several examples, we found that framing manipulations impacted the pattern of response on the discounting and demand tasks used. Precisely, framing of high risk social activities increased sensitivity to risk for social distancing, framing delay as a delay to result increased sensitivity to delay for test procurement, and framing vaccine safety in a negative valence increased sensitivity to efficacy (thereby more steeply reducing vaccine acceptance). The use of simulated discounting and demand tasks, furthermore, provided a substantive benefit over traditional single discrete-choice forms of assessment (e.g., “Would you get a COVID-19 test?). Such single discrete-choice methods fail to isolate and control for factors that may contribute to differences observed between and within-people (e.g., differences in hypothesized delays, risk, efficacy, or safety). Responding under such methods may therefore be attributable to any of these uncontrolled factors with differing implications for public policy based on the specific mechanism(s) impacted.

These contributions should be considered within the limitations of this study. For one, we restricted sampling to a crowdsourced platform. An extensive body of literature suggests the reliability and validity of data collected through crowdsourced platforms is favorable in comparisons to other convenience methods like undergraduate student pools [64, 65]. Nevertheless, crowdsourcing approaches are still convenience sampling and present some bias such that sampling favors towards younger participants [64]. Crowdsourcing in this context served as an ideal data collection method for generating a large and geographically diverse sample in the face of a rapidly changing public health context in which in-person study was challenging, if not impossible, for this purpose. Some tasks were also evaluated in the same sample of participants as noted for each analysis throughout. A relatively high number of participants displayed non-systematic responding, which may be related to the use of a comparably low prior task approval rate and/or the use of a one-step rather than two-step (i.e., screener and survey) sampling approach [66-68]. Relevant to the specific contributions of these data for COVID-19 and related pandemic responses, our findings are potentially limited by the use of a between-subject manipulation, specific features of the vignette, and collection at a single point in time. Decisions on what was a between- and within-subject manipulation came after careful consideration to maximize a preference for within-subject designs while recognizing design options likely to result in substantive carry-over bias. These findings are also limited to the hypothetical scenarios used and it is likely that variations of these scenarios would produce further variations in behavior [69-71]. Although the tasks presented were hypothetical in nature, extensive work have found hypothetical versions of these tasks are a reasonable proxy for procedures using real consequences [32, 72-75]. The flexibility of these procedures and ability to evaluate hypothetical decision-making for which incentivized responding is either unpractical or unethical is a major strength insofar as they afford the opportunity to evaluate and compare in short succession a variety of potential contexts relevant to public health response.

The COVID-19 epidemic has challenged a spectrum of sciences to reconsider their ability to quickly translate methods to understand, model, and mitigate contagion. The field of behavioral and decision-making science has a rich and productive history addressing issues of societal importance including disease prevention and health promotion. Behavioral economics is, perhaps, a prime aspect of how behavioral science can leverage its methods toward this end, given its ability to address difficult-to-measure behavior and quantify outcomes that are readily translatable to public health researchers and officials. Here we show how merging conceptual ideas from a cognitive and operant psychology behavioral economics using both discounting and demand methods to provide novel understanding to behavioral components of a global pandemic (COVID-19). Ultimately, these data provide an example of the adaptability and translational utility of behavioral economics when current and future public health crises necessitate behavioral insight and solutions.

## Data Availability

Limited data set and code to replicate primary analyses is available at https://osf.io/wdnmx/?view_only=37323f9431aa4c91a0e7209054058dbe.

