## Supplemental Materials for "Integrating Operant and Cognitive Behavioral Economics to Inform Infectious Disease Response: Prevention, Testing, and Vaccination in the COVID-19 Pandemic"

**Experiment 1 Vignettes**

*(bracketed italics included for participants randomized to the label condition)

**Mild Symptom Vignette**

For the next set of questions, we ask you to imagine you have plans to attend a large social event (e.g., a sporting event, concert, lecture, theatrical performance, gala/reception) with over 200 people who will be seated in close proximity to one another. You have already spent $50 for admission and promised several of your friends you would be attending. The event is being held in a community with a population of approximately 100,000 people.

The day before the event you learn there is chance that someone in the community, who may be at the event or have been in contact with others at the event, exhibits the following **FIVE (5)** symptoms [*(this group of symptoms is classified as* ***mild****)*]:

- Dry cough
- Fatigue
- Fever
- Shortness of breath
- Headache

**Severe Symptom Vignette**

For the next set of questions, we ask you to imagine you have plans to attend a large social event (e.g., a sporting event, concert, lecture, theatrical performance, gala/reception) with over 200 people who will be seated in close proximity to one another. You have already spent $50 for admission and promised several of your friends you would be attending. The event is being held in a community with a population of approximately 100,000 people.

The day before the event you learn there is chance that someone in the community, who may be at the event or have been in contact with others at the event, exhibits the following **SIX (6)** symptoms [*(this group of symptoms is classified as* ***severe****)*]:

- Dry cough
- Fatigue
- Fever
- Shortness of breath
- Headache
- Difficulty breathing (requires a medical ventilator)

**Response Options**

Please rate your likelihood of attending that event at each % chance someone in the community is presenting those symptoms (note: you must click on each slider to register a response):

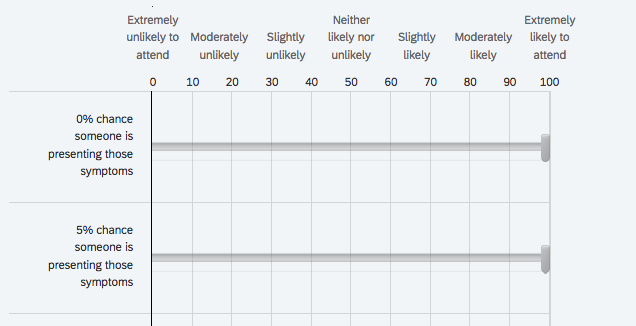

**Experiment 2 Vignettes**

*activities were personalized through pipped text

**Activity Vignette**

Imagine that the state you live in is currently open for social events. You just made plans to
 ${q://QID32/ChoiceGroup/SelectedChoices}.

**Label Manipulation**

According to health authorities in your area, this activity is of **[Low-to-Moderate Risk**/**High Risk]** when assuming you and those around you follow the recommended safety protocols.

**Response Options**

Please rate your likelihood of attending that event at each % chance someone at that event may have symptoms related to COVID-19 (e.g., a dry cough):

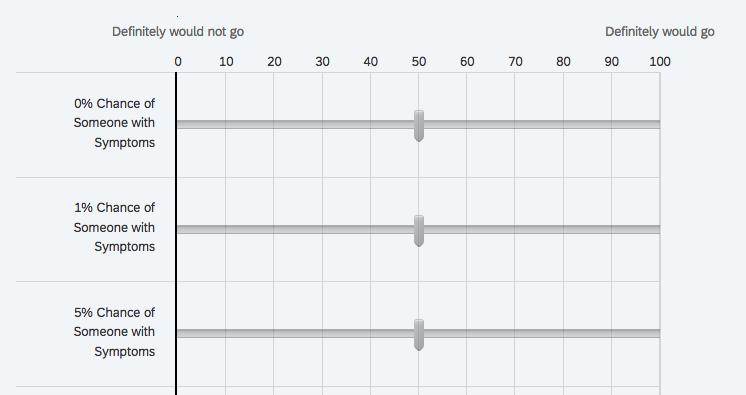

**Experiment 3 Vignettes**

**Preamble**

Think about what you would do in following situation for the people on your list of 100 people closest to you in the world ranging from your dearest friend or relative at position #1 to a mere acquaintance at #100.

**Asymptomatic Vignette**

Imagine that a week ago, you were with a large crowd of people (~50 people) for about an hour.

You are now going to meet with one of the people on your list of 100. You are going to be in a situation where you will **not** be able to maintain social distance (6 feet) the whole time you are with them.

**Symptomatic with No Positive Test Vignette**

Imagine that a week ago, you were with a large crowd of people (~50 people) for about an hour. Over the past week, you've developed a **dry cough and low grade fever, but otherwise feel okay**.

You are now going to meet with one of the people on your list of 100. You are going to be in a situation where you will **not** be able to maintain social distance (6 feet) the whole time you are with them.

**Symptomatic with Positive Test Vignette**

Imagine that a week ago, you were with a large crowd of people (~50 people) for about an hour. Over the past week, you've developed a **dry cough and low grade fever, but otherwise feel okay. However, you got a COVID test as a precaution and just found out that you tested positive.**

You are now going to meet with one of the people on your list of 100. You are going to be in a situation where you will **not** be able to maintain social distance (6 feet) the whole time you are with them.

**Response Options**

For each of the following people, please rate the likelihood that you would wear a face mask in that meeting.

Please select the likelihood you would wear a face mask if meeting up with each of the following people given the conditions above:

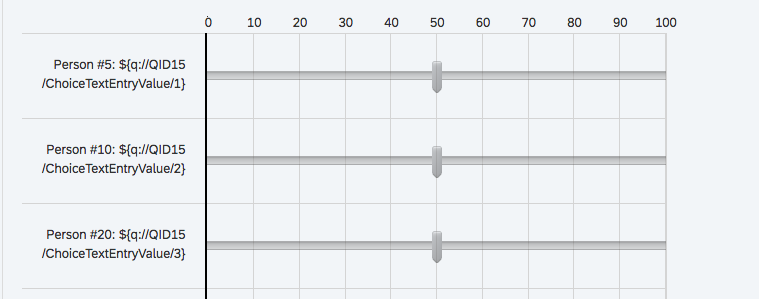

* piped text was used to personalize responses

**Experiment 4 Vignette**

For the next set of questions, we ask you to imagine you have attended a large social event (e.g., a sporting event, concert, lecture, theatrical performance, gala/reception) with over 200 people who were seated in close proximity to one another. One week later, you begin exhibiting the following symptoms:

- Cough
- Fever
- Shortness of breath

Assumptions:

- At least one other person in your county has tested positive for an infection.
- Your nearest hospital/clinic has a testing kit available to diagnose whether you have an infection.
- There are no other diagnostic testing kits in your area.
- The diagnostic testing kit is approved by the Centers for Disease Control and Prevention (CDC).
- You have the same income and savings as you do right now.

What is the likelihood you would pursue testing at your nearest hospital/clinic for an infection if your out-of-pocket cost (that is, not covered by insurance) for the diagnostic testing was (note: you must click on each slider to register a response):

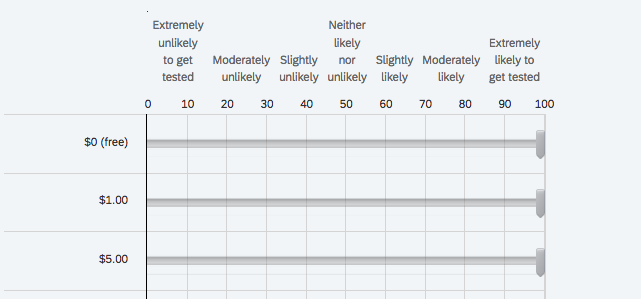

**Experiment 5 Vignettes**

*(bracketed are manipulations based on cost condition)

**Delay to Receive Vignette**

Imagine a situation in which test kits (nasal swabs) were available for **[free/$125]** via **home delivery with a shipping delay** for any citizen who wanted to get tested. These tests are easy to use and take less than 5 minutes to be administered, and **only 15 minutes to receive your results**. Results of these tests would allow individuals to know if they have COVID-19.

The following questions ask about whether or not you would get a COVID-19 swab test for **[free/$125]**, given a series of **delays to receiving the test itself**.

Assumptions:

- The swab test is easily administered and can be done with minimal assistance
- [The swab test is available to you without cost (free) OR The swab test is available to you at a one-time, out-of-pocket expense of $125. You will not be reimbursed for this payment.]
- You have the same income/savings as you do now
- You have no access to any other tests available for COVID
- The test must be administered at the time of receiving it (you can’t save the test kit to use at a later date)
- This test must only be used for you (you cannot use this test for friends or family members)
- You will receive the test at your home, after a shipping delay
- You will receive your results 15 minutes after taking the test
- This test is 100% accurate

Remember that there are no “right” or “wrong” answers. Please respond honestly, as if you were actually in this scenario.

Given the above scenario, please **indicate whether or not** you would get a COVID-19 swab test for **[FREE/$125]**given the **delay to receiving the test itself**.

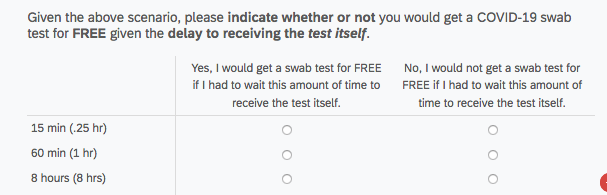

**Delay to Results Vignette**

Imagine a situation in which test kits (nasal swabs) were available for **[free/$125]** via **same-day home delivery** for any citizen who wanted to get tested. These tests are easy to use and take less than 5 minutes to be administered, but **you have to wait for the results**. Results of these tests would allow individuals to know if they have COVID-19.

The following questions ask about whether or not you would get a COVID-19 swab test for **free**, given a series of **delays to receiving results**.

Assumptions:

- The swab test is easily administered and can be done with minimal assistance
- [The swab test is available to you without cost (free) OR The swab test is available to you at a one-time, out-of-pocket expense of $125. You will not be reimbursed for this payment.]
- You have the same income/savings as you do now
- You have no access to any other tests available for COVID
- The test must be administered at the time of receiving it (you can’t save the test kit to use at a later date)
- This test must only be used for you (you cannot use this test for friends or family members)
- You will receive the test at your home, the same the day you request it
- You will wait to receive the results after a delay
- This test is 100% accurate

Remember that there are no “right” or “wrong” answers. Please respond honestly, as if you were actually in this scenario.

Given the above scenario, please **indicate whether or not** you would get a COVID-19 swab test for **[FREE/$125]** given the **delay to receiving the results**.

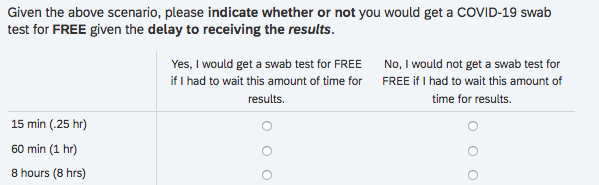

**Experiment 6 Vignettes**

**COVID-19 Vaccine Vignette**

Imagine that you went to a doctor to get a seasonal flu vaccine. As a part of getting the flu vaccine, you **can also get a COVID-19 vaccine at no charge.**

Assumptions:

- The vaccine is available to you without cost (free)
- You have no access to any other vaccines available for COVID-19
- These vaccines are approved by the FDA

**Flu Vaccine Vignette**
Imagine that you went to a doctor to get a COVID-19 vaccine. As a part of getting the COVID-19 vaccine, you **can also get a seasonal flu vaccine at no charge.**

Assumptions:

- The vaccine is available to you without cost (free)
- You have no access to any other vaccines available for the flu
- These vaccines are approved by the FDA

**Response Options**

Given the above scenario, please **indicate whether or not** you would you also get the flu vaccine if it reduced chances of flu symptoms by each of the following percentages (X%):

**Opt-Out**

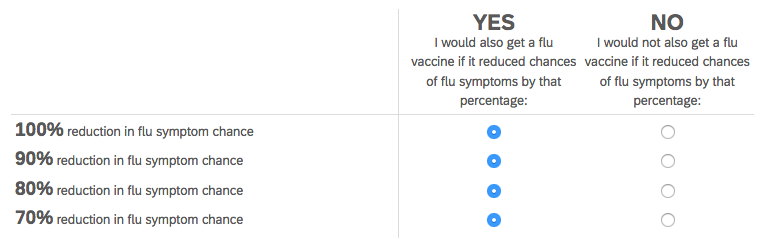

**Opt-In**
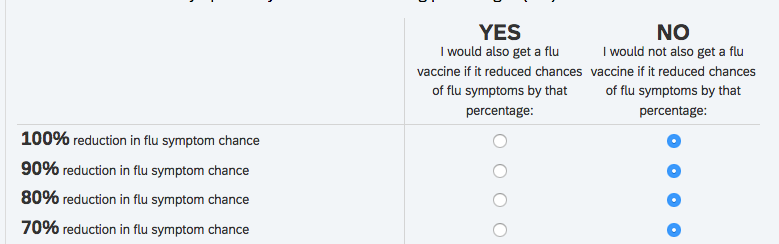

**Open Response**

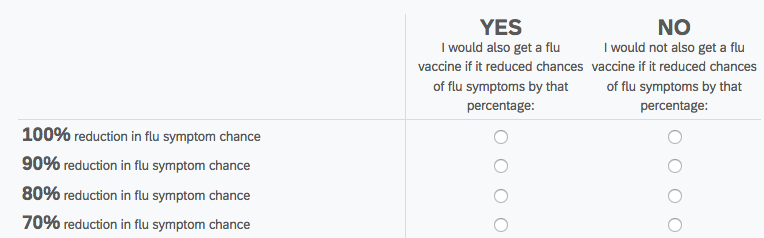

**Experiment 7 Vignettes**

**7 Month Development Framing**

Suppose a COVID-19 vaccine was **developed in a total of 7 months**, beginning in late March, 2020 with **delivery to the general population by late October, 2020**. Imagine the vaccine has been **approved by the Food and Drug Administration** (FDA). Importantly, **[5% of the scientific community declares the vaccine unsafe/95% of the scientific community declares the vaccine safe]**. You can get the vaccine through a doctor, at **no cost** to you.  

Assumptions:

- The vaccine is easily administered
- The vaccine is available to you without cost (free)
- You have no access to any other vaccines available for COVID
- The vaccine must be administered at the time of receiving it (you can’t save it to use at a later date)
- This vaccine must only be used for you (you cannot use this vaccine for friends or family members)
- This vaccine is approved by the FDA
- [5% of the scientific community declares the vaccine unsafe/95% of the scientific community declares the vaccine safe]

**12 Month Development Framing**

Suppose a COVID-19 vaccine was **developed in a total of 12 months**, beginning in late March, 2020 with **delivery to the general population by late March, 2021**. Imagine the vaccine has been **approved by the Food and Drug Administration** (FDA). Importantly, **[5% of the scientific community declares the vaccine unsafe/95% of the scientific community declares the vaccine safe]**. You can get the vaccine through a doctor, at **no cost** to you.  

Assumptions:

- The vaccine is easily administered
- The vaccine is available to you without cost (free)
- You have no access to any other vaccines available for COVID
- The vaccine must be administered at the time of receiving it (you can’t save it to use at a later date)
- This vaccine must only be used for you (you cannot use this vaccine for friends or family members)
- This vaccine is approved by the FDA
- [5% of the scientific community declares the vaccine unsafe/95% of the scientific community declares the vaccine safe]

**Response Options**

Remember that there are no “right” or “wrong” answers. Please respond honestly, as if you were actually in this scenario.

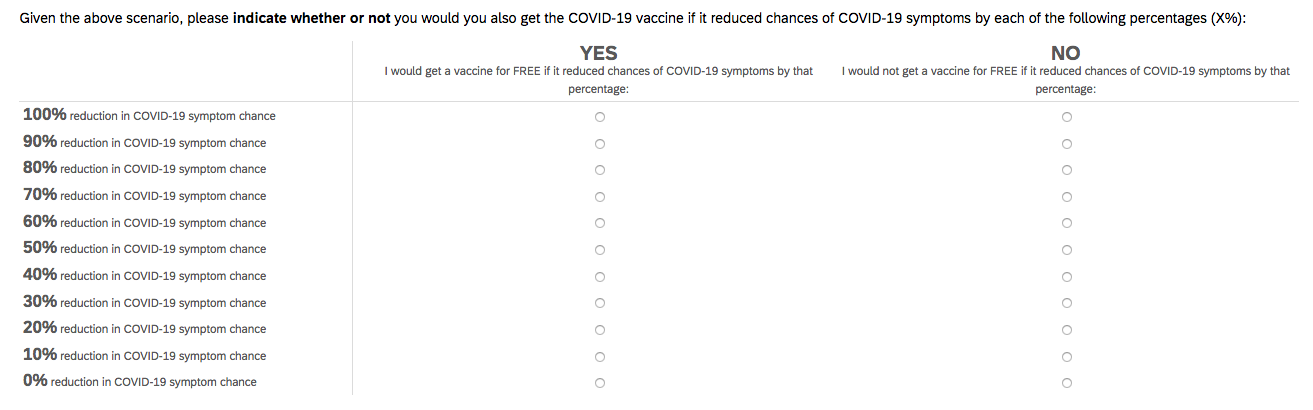
